# Medical Devises Regulation in Zimbabwe: An Evaluation of operational readiness

**DOI:** 10.1101/2023.06.08.23291162

**Authors:** Charles Chiku, Talkmore Maruta, Fredrick Mbiba, Justen Manasa

## Abstract

**Background:** Regulation of medical devices has seriously lagged, especially in Resource-Limited Settings (RLS). There are disparities in regulating medical devices; in the African region, it is below the global average. This may translate into poor access to quality-assured medical devices and result in undesirable health outcomes. Operational readiness to regulate medical devices in Zimbabwe at the Medicines Control Authority of Zimbabwe (MCAZ), the designated National Regulatory Authority (NRA), is vital for planning and implementation. The study aimed to assess the readiness of the MCAZ to regulate medical devices by identifying the strengths and gaps and proposing an institutional development plan that can be monitored and evaluated to assess progress over time.

**Methods:** Quantitative study was conducted using the World Health Organization (WHO) Global Benchmarking Tool+ medical devices (GBT+ medical devices) methodology to evaluate the medical devices regulatory oversight at the MCAZ. Data were collected between June and August 2022 using standard checklists to assess the quality of implementation of medical devices’ regulatory functions; National Regulatory System (RS), Registration and Market Authorisation (MA), Vigilance (VL), Market Surveillance and Control (MC), Licensing Establishment (LI), Regulatory Inspection (RI), Laboratory Testing(LT), and Clinical Trials (CTs) Oversight.

**Results:** The MCAZ attained maturity level 1, with an RS score of 79%, MA-44%, VL-27%, MC-40%, LI-62%, RI-68%, LT-88%, and CT-18%. Condoms and gloves were the only regulated medical devices. Indicators on legal provisions, regulations, and guidelines across the regulatory functions were below the optimum requirement.

**Conclusion:** The legal provisions, regulations, and guidelines are inadequate for effectively regulating medical devices. The medical devices regulation requires review for it to be robust and fit-for-purpose, responsive, oriented to the outcome, predictable based on a standard and transparent approach, and the level of scrutiny proportionate to the risk classification of the medical device.

## Introduction

Medical devices are essential for the public to reach the highest health standards. Although regulations for medicines and vaccines have existed for many years, regulation of medical devices, including In Vitro Diagnostics (IVDs) medical devices, has seriously lagged, especially in resource-limited settings (RLS). A medical device is “any instrument, apparatus, machine, appliance, implant, reagent for in vitro use, software, material, or related article used for a specific medical purpose” (1,2). An IVD medical device is used to examine human specimens to provide diagnostic information, monitoring, or compatibility purposes (3).

### Medical Device Regulations

Medical Device Regulations are a set of laws and regulations governing clinical trials, manufacturing, and distribution of medical devices to ensure they are safe and perform as intended by their manufacturers (4,5). The primary objective of regulating medical devices is to facilitate access to safe, effective medical devices with acceptable performance and quality to ensure safety for patients and users.

Regulatory processes in the African region are not well documented compared to the US Food and Drug Administration (FDA) and the European Union’s European Medicines Agency (6). However, despite the regulatory processes being well documented in the US and the EU, they have not been immune to challenges associated with product recalls, adverse incident reports and removing some medical devices from the market (7,8). These systems are not easily adoptable by RLS due to their prohibitively high maintenance costs and the capacity required to ensure continuous improvement. Harmonising regulatory systems across different settings is essential to reduce the regulatory burden on manufacturers and other economic operators in getting their products on the market. It has also been concluded that poor regulatory systems in developing countries make it difficult for manufacturers to introduce their products in these markets, resulting in limited access to them (9,10).

In general, there are mechanisms to reduce the regulatory burden on manufacturers and the workload on the regulators. These include convergence, harmonisation, reliance and recognition. Convergence is an approach that seeks to make regulatory requirements similar across countries and adopt internationally recognised standards, technical guidance documents, shared scientific principles and procedures to ensure that public health is protected considering the local context. It does not necessarily mean the harmonisation of laws. Harmonisation is the process by which technical guidelines are developed to be uniform across participating authorities. Reliance is the act whereby the regulatory authority in one jurisdiction takes into account and bases its decision on an evaluation conducted by another regulatory authority. In contrast, the relying authority remains responsible for its decisions. The reliance mechanism is meant to reduce the barriers to access of IVDs due to unclear regulations and a lack of legal provisions to implement it. Regulatory reliance enables healthcare systems to;

- accelerate global access to safe and quality health technology,
- increase efficient use of resources and avoid duplication of efforts,
- reduce uncertainties for innovators and improve harmonisation in regulation,
- promote more consistent and robust responses to crises (2).

Recognition is a mechanism that allows approval from one regulatory authority to be given equivalent weight in another jurisdiction. Recognition indicates that evidence of conformity with the regulatory requirements of country X is sufficient to meet country Y(11).

### Disparities in Medical Device Regulations

There are disparities in the availability of medical device regulatory services between the global and WHO Africa (Afro) region. About 40% of countries in the WHO Afro region had no regulations for medical devices, 32% had some regulations, and 28% had no regulations.

In contrast, at the global level, 58% of all WHO member states indicated that they had regulations on medical devices. Zimbabwe was presented with the element of only placing medical devices on the market. Premarket and post-market elements were unavailable according to the survey published in 2016 by the WHO(12). This gap in medical device regulation between the Afro region and the global average is crucial as it may translate to lower-quality medical devices and limited patient access to healthcare technologies. The lack of harmonised regulations may result in a regulatory burden for manufacturers to introduce their products on the market. The fundamental characteristics of a robust and fit-for-purpose regulatory system are responsiveness, oriented to the outcome, predictability based on a standard and transparent approach, and the level of scrutiny to assess the conformity of the medical devices to set requirements must be proportionate to the risk classification of the medical device. The regulatory system and institutions must be independent (13).

### Medical Device, including IVD Regulations in Zimbabwe

There is a dearth of medical device literature specific to Zimbabwe. Hubner et al. conducted a study to determine the evolving landscape of medical device regulations in member countries of the Surgeons of Central, Eastern and Southern Africa (COSECA). Zimbabwe is a member of COSECA. The study used a systematic review of the literature on medical devices. It was concluded that Zimbabwe had a legal framework for regulating medical devices, conformity assessment (evaluation conducted to approve medical devices to be granted market access), and import and export and post-market surveillance for condoms and gloves only (9). Contrary to the finding, the WHO survey concluded that Zimbabwe only had elements of placing medical devices on the market without premarket and post-market elements. It is unclear whether the framework in Zimbabwe has evolved and is robust enough to effectively regulate medical devices, including IVDs.

Another study conducted by Mwedzi et al. in monitoring the progress in regulatory systems strengthening in the Southern African Development Community (SADC)-Zimbabwe is a member state of SADC. It was determined that the NRAs with a normative legal framework for medical devices were just above 50%, and less than 50% of the member states had a legal framework for regulating IVD medical devices or imaging equipment. The study looked at premarket approval, to a lesser extent, clinical testing and post-marketing surveillance (13). Furthermore, the results lacked granularity since they were anonymised and unlinked, making it difficult to conclude the regulatory status of Zimbabwe.

Similarly, Kniazkov et al. surveyed to map existing frameworks, mechanisms and approaches to prevention, detection and response (PDR) to Sub-Standard and Falsified (SF) medical products. Findings pointed to deficiencies in policies and implementation plans despite most countries having the mandate and legislation to deal with substandard and falsified (SF) medical products. This study also lacked specific results for Zimbabwe due to unlinking and anonymising the countries. Moreover, the study was for medical products in general without specifying medical devices (14).

A qualitative study to assess the regulation of HIV-Self Testing IVDs in Malawi, Zambia and Zimbabwe was conducted by Dacombe et al. The study aimed to document the emerging regulatory landscape and perceptions of key stakeholders involved in HIVST policy and regulation before implementation in three low- and middle-income countries. It was found that the reference laboratory monitored the quality and performance of HIV-Self Testing IVDs used in the public sector in all three countries. However, the mandate to regulate HIV-Self Testing IVDs overlapped between the Medical Laboratory and Clinical Scientists Council of Zimbabwe and the Medicines Control Authority of Zimbabwe (MCAZ). Stakeholders indicated they had a poor understanding of the process and requirements for HIVST regulation and a lack of clarity and coordination between organisational roles (15).

The extent to which Zimbabwe’s regulatory system governance is an essential driver for implementation has yet to be adequately appraised due to a lack of studies specific to the Zimbabwean context. Recent studies have shown an absence of medical device regulation literature in the COSECA region. The same study concluded that Zimbabwe had no formal regulatory system for medical devices except for gloves and condoms (9). The study also concluded that MCAZ is a regulatory authority mandated to regulate medical devices. However, another study examining HIV Self-Testing IVDs in Zambia, Malawi and Zimbabwe concluded that it was unclear which institution was mandated to regulate IVDs in Zimbabwe (15).

Our study aimed to unpack the landscape of medical device regulation in Zimbabwe, focusing on the MCAZ as the NRA using the GBT + medical devices. The GBT tool was created after harmonising benchmarking tools for health products. The harmonisation was conducted to maximise outcomes and reduce the regulatory burden for the NRAs and manufacturers. The GBT is “a game changer in strengthening national regulatory capacity” through a global standardised manner of evaluating regulatory systems (16). An effective regulatory system is essential to strengthening health and improving health outcomes. Better health outcomes for the population are achieved through access to safe medical devices of acceptable quality, performance and effectiveness. This is the goal of effective medical device regulation.

On the one hand, due to SF medical devices and associated economic costs, underdeveloped regulatory systems may severely affect public health. Underdeveloped regulatory systems are associated with the regulatory burden for manufacturers to comply with unclear or different regulations when seeking premarket approvals in other countries. Therefore, some manufacturers may be reluctant to introduce their medical devices in countries with underdeveloped regulatory systems, which makes it difficult for the population to access much-needed medical devices. Conversely, the respective NRAs must provide balance by ensuring public health and users’ protection through approved, safe, effective, acceptable quality and performance medical devices (10). The readiness of the MCAZ to effectively regulate medical devices beyond condoms and gloves is unknown. MCAZ is a statutory body established by Parliament, The Medicines and Allied Substances Control Act (MASCA) [Chapter 15.03], to regulate medicines and Allied Substances. Therefore, benchmarking the MCAZ is crucial in assessing its readiness to regulate medical devices effectively (17).

The study is significant because Zimbabwe is expanding the regulatory system by expanding the scope of health products regulated to include IVDs. Fundamentally, independent and objective benchmarking be conducted to identify strengths and deficiencies. The benchmarking needs to be standardised according to globally acceptable standards. The WHO GBT is one of the globally acceptable ways of benchmarking the maturity level of NRAs (18). The tool uses a set of indicators to evaluate the NRA. It uses it as input in developing a road map to guide establishing, implementing and maintaining an effective medical devices regulatory system. Additionally, the road map works as a tool for the regulatory system to be responsive, predictable, transparent and proportional to the public health risks will ensure the population access to safe and acceptable quality and performance medical devices.

## Methods

### Study design

We conducted a quantitative study using the WHO GBT+ medical devices methodology to evaluate the medical devices regulatory oversight at the MCAZ in Zimbabwe between June and August 2022. The following regulatory functions were assessed following the medical devices’ life cycle:

- National Regulatory System (RS)-The legal and regulatory framework supports the regulatory system’s functions to ensure the quality, safety, and performance of medical products.
- Registration and Market Authorisation (MA)-The issuance of marketing authorisations (also referred to as product licensing or registration) when medical products have met the requirements of standardised conformity assessment.
- Vigilance (VL)-The science and activities relating to the prevention, detection, assessment, and understanding of adverse effects or any other medical product-related problems for guaranteeing that medical product continues to meet quality, safety and performance requirements throughout the product’s lifecycle.
- Market Surveillance and Control (MC)-The function of assuring ongoing compliance of the products placed on the market with quality, safety and performance requirements.
- Licensing Establishment (LE) – The function of guaranteeing the quality, safety, and performance of medical products used within or exported out of the country through licensing of establishments involved in the value chain and life cycle of the medical products.
- Regulatory Inspection (RI)-Auditing establishments throughout the value chain and life cycle of medical devices to ensure compliance of these establishments with laws, regulations, approved standards, norms, and guidelines.
- Laboratory Testing(LT)-The independent performance verification of the manufacturer’s performance claims by the NRA to support premarket approval or a variation or change to marketing authorisation.
- Clinical Trials (CTs) Oversight-Refers to the legal mandate of the NRA to authorise, regulate and, if necessary, terminate CTs.

The regulatory functions were assessed using a set of indicators that were divided into the following nine categories:

1. Legal provisions, regulations and guidelines.
2. Organisation and governance.
3. Policy and strategic planning.
4. Leadership and crisis management.
5. Transparency, accountability and communication.
6. Quality and risk management systems.
7. Regulatory process.
8. Resources (including Human, financial, infrastructure, equipment and information management systems).
9. Monitoring progress and assessing impact.

Sub-indicators are grouped under a parent indicator to compile overall scores (19).

### Study setting

The benchmarking of the MCAZ was conducted between June and August 2022 since the institution is a statutory body established to regulate medicines and allied substances according to the Medicines and Allied Substances Control Act (MASCA) [Chapter 15.03].

### Data Analysis

Descriptive analysis was performed to describe the quality of regulatory function implementation based on the cumulative score of sub-indicators. The Computerised GBT (cGBT) scoring algorithm was used to determine the implementation status of each indicator for each regulatory function. Scoring was done as a measure of assessing the quality of the implementation of each sub-indicator as follows:

1. Not implemented (NI): no evidence was provided to demonstrate any degree of implementation of the sub-indicator. This status was assigned where a score of 0% as a percentage is attained.
2. Ongoing implementation (OI): some actions/steps/activities were taken towards implementing the concerned sub-indicator. However, the sub-indicator still needs to be implemented in full. A score of 25% was assigned to this category (a score of 25% as a percentage).
3. Partially implemented (PI): some actions/activities showed the full implementation of the sub-indicator; however, such full performance is recent or relatively new, with little cumulative data for consistent execution. Supporting documented evidence was expected to show the current full implementation of the concerned sub-indicator. For mathematical scoring, ‘partially implemented’ is scored as 0.75 out of one (i.e., 75% as a percentage).
4. Fully implemented (I): some actions/activities demonstrate the consistent and full implementation of the sub-indicator over time. Supporting evidence is expected to illustrate the full, consistent implementation of the sub-indicator (i.e., shown over time and through repetition of the process and outcome). ‘Fully implemented’ is scored as one out of one (i.e., 100% as a percentage).

The cGBT algorithm was used to assign maturity levels (MLs) based on the cumulative scoring of the sub-indicators under that function. For a regulatory function to reach a specific ML, a specified percentage of sub-indicators must be scored as ‘fully implemented’ (19).

### Ethical Considerations

Ethical approval for this study was obtained from the Medical Research Council of Zimbabwe (MRCZ/A/2900). Participation was voluntary. Participants were able to stop the interview at any time without explanation. Written informed consent was obtained from each study participant before each interview. The interview content and the interviewee’s identity were kept anonymous.

## Results

The regulatory functions that were assessed were all at maturity level 1. Maturity level 1 is the minor level to measure each indicator’s implementation quality. Figure 1 shows the maturity level of each regulatory function. The Lot Release regulatory function was not assessed because it does not apply to medical devices.

**Figure 1.**
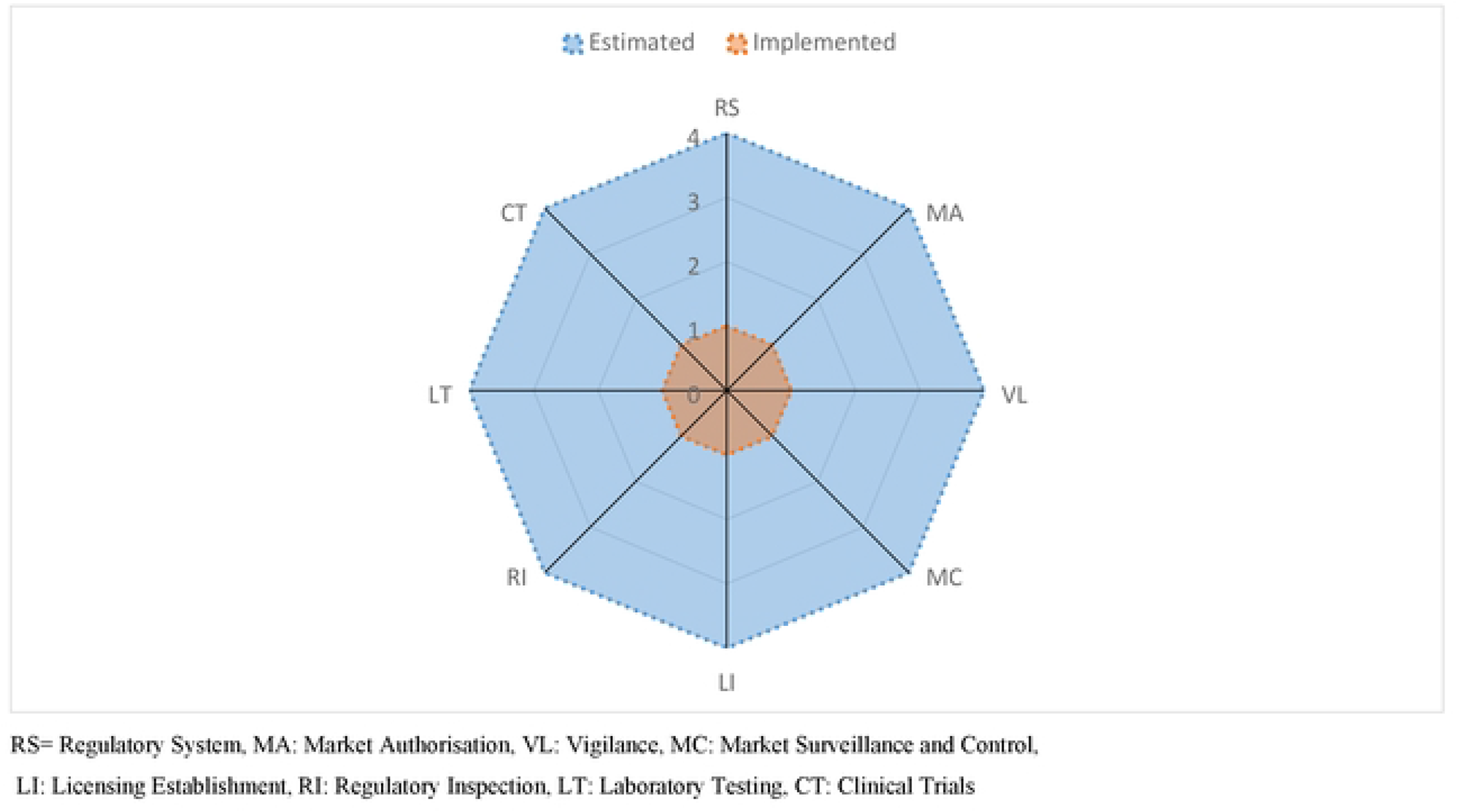
The Maturity Level for the Regulatory Functions assessed at MCAZ between June and August 2022.

The quality of implementation of each regulatory function ranged between 18% to 88%. Clinical Trial was the least, with 18%, and Laboratory Testing scored 88%. Figure 2 shows the performance of each regulatory function that was assessed.

**Figure 2:**
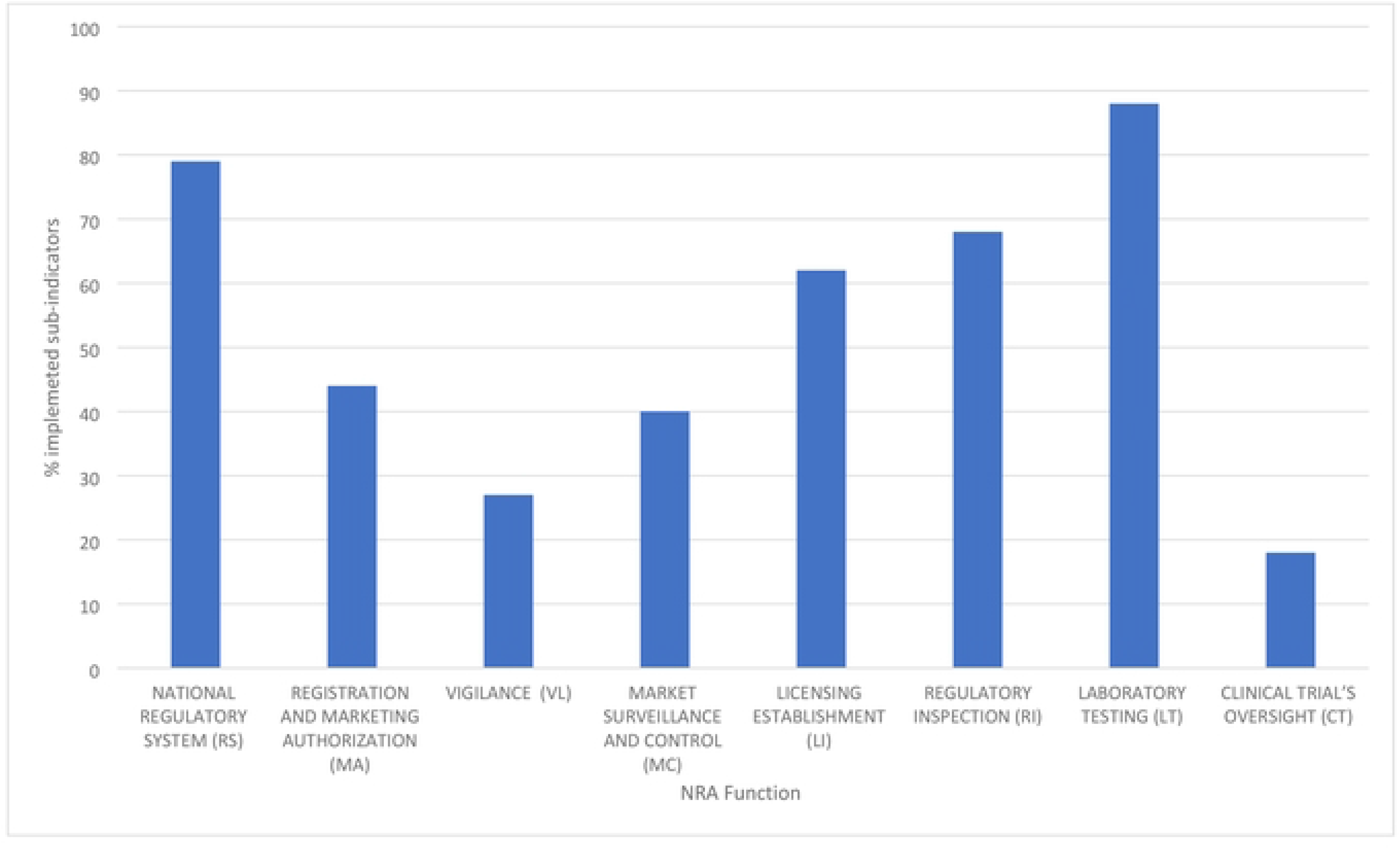
Quality of Implementation of regulatory functions at MCAZ between June and August 2022.

The national regulatory system function did not have explicit legal provisions and regulations that required medical devices to meet specific safety, quality and performance requirements throughout their lifecycle. Additionally, the MASCA did not define the roles and responsibilities of institutions involved in the medical devices regulatory system. Similarly, there were no legal provisions and relevant regulations to take action on the recall, suspension, withdrawal and destruction of SF medical devices.

MA function fell short in legal provisions and regulations that mandate medical devices to be assessed against stipulated requirements before accessing the market. No requirements require medical devices to be subjected to a conformity assessment proportional to the medical device’s risk class. Draft IVDs regulations were in place pending approval for implementation.

The VL function was not robust enough to prevent, detect, assess, and understand the adverse effects or any other medical device-related problems throughout the medical devices’ lifecycle, except for condoms and gloves.

Efforts were ongoing to address market surveillance and controls. There were no legal provisions and regulations to control import activities. However, Import and Export regulations for medical devices were in draft format, pending approval. Legal provisions and regulations authorise market surveillance and control activities, including product sampling from different supply chain points, were unavailable. The NRA did not have legal provisions and regulations to address its role in dealing with SF medical devices.

No legal provisions and regulations required economic operators involved throughout the value chain of medical devices to be registered based on Good Practices (GXP) compliance. Furthermore, there were no legal provisions to empower the NRA to issue, suspend or revoke licenses for establishments for medical devices besides condoms and gloves.

No legal provisions, regulations or guidelines were required to define the regulatory framework of inspection and enforcement for medical devices, except for condoms and gloves. Additionally, there were no updated national GXP regulations, norms or guidelines that are mandatory to guide economic operators on their submissions.

The laboratory testing function was well established and implemented for condoms and gloves. Although the legal basis is unclear, demonstrable steps have been taken towards establishing one for IVD medical devices. In addition, there is no evidence of legal provisions or regulations allowing reliance on and recognition of other laboratories’ regulatory decisions.

There were no legal provisions or regulations for CT oversight. Moreover, no legal provisions or regulations required notification to and authorisation from the NRA diversions from the original protocol of the CTs. Lastly, there was no evidence of legal provisions, regulations or guidelines requiring that investigational medical products (IMPs) comply with good manufacturing practices for IMPs.

Cross-cutting indicators were generally in place; policy and strategic planning, leadership and crisis management, transparency, accountability and communication, quality and risk management systems, regulatory process, resources (Human, financial, infrastructure, equipment and information management systems), monitoring progress and assessing impact. However, the indicators were sufficient for regulating condoms and gloves only. There were some vacant staff positions compared to the current establishment of the NRA. The competence framework was sufficient for the existing scope of regulated medical devices (condoms and gloves) but not for other medical devices. Transparency and accountability needed to be improved as the NRA was not publishing public reports, excerpts of regulatory assessments conducted (market authorisation, laboratory or inspection reports), and lists of all approved or rejected applications.

## Discussion

### RS

Medical devices, including IVDs in Zimbabwe, are not adequately regulated due to the low score of less than 80%. The ineffective regulation of these health products is because of an underdeveloped legislative framework. The MASCA was promulgated specifically to regulate medicines and Allied Substances. However, the term “Allied Substances” was not defined, leaving a gap in the scope of products that can be regulated under this legislation. Hubner et al. concluded that the Zimbabwean legislation did not define medical devices (9). Therefore, our finding is consistent with Hubner et al.’s finding.

The MCAZ seems not to have a clear mandate to regulate medical devices. The implications are the non-regulation of medical devices because of the undefined regulatory framework for the national regulatory system. Considering no standard definition of medical devices and the lack of a well-defined regulatory framework may have resulted in the unavailability of essential principles of safety and performance and risk-based classification of medical devices, as concluded by Hubner et al. Overall, the maturity level of the regulatory system was found to be ML 1, which is consistent with the conclusion by Hubner et al. that Zimbabwe did not have a formal regulatory process for medical devices, except for some aspects to regulate condoms and gloves (9).

The findings clearly show that Zimbabwe’s medical devices regulatory system has not been responsive, outcome-oriented, predictable, risk proportionate to public health risk and independent; these are critical characteristics of a robust regulatory system. The finding agrees with what Dube-Mwedzi et al. said about efforts to strengthen regulatory frameworks for medicines, whilst focus on medical devices has been lower (13). Yet, poor-quality medical devices significantly impact healthcare outcomes. It is widely accepted now that the value of medication rests mainly on the accuracy of the diagnosis. Thus, using poor-quality medical devices and diagnostics undermines the effectiveness of efforts to make good quality, safe and effective medicines available. Investing more efforts into strengthening frameworks for medical devices could be recommended as a priority for the SADC countries. Dube-Mwedzi also stressed the need to strengthen governance to make the national regulatory authority more efficient. Looking at the Zimbabwean context, following an institutional approach that involves implementing the “best practices” seems plausible to harmonise the regulations. The other option requires experimentation and prioritisation of country-specific challenges, which may be challenging and costly for countries without experience. The problem-driven approach diverges from the institutional approach by prioritising country-specific issues and enforcement over the blanket implementation of “best practices.” This approach allows for feedback loops and greater policy experimentation as problems arise (20). Therefore, the MCAZ must be empowered to regulate medical devices based on a solid legal foundation. The methodology in our study differed from Dube-Mwedzi et al.’s because the latter focused on the SADC member states, and the data presented needed to be more granular to identify where Zimbabwe stood in the study. In our study, we had an in-depth look at the Zimbabwean context.

Furthermore, the methodology in this study looked at registration and licensing (import and export controls), clinical testing and PMS regulatory functions. In addition to the stated regulatory functions, the study conducted by this researcher looked further at indicators specific to regulatory systems, market authorisation, Licensing establishment, product performance evaluation, and regulatory inspection and quality management system audits. The findings on the additional regulatory functions are stated below.

### MA

The quality of performance for this regulatory function was 44% due to the lack of legal provisions and regulations to mandate the registration of medical devices before accessing the market. The legal provisions, regulations and guidelines for medical devices should also support notification/listing low-risk products. The International Medical Devices Regulatory Framework (IMDRF) and its predecessor, the Global Harmonisation Working Party (GHTF), recommend that medical devices be assigned to one of four classes based on a set of rules. This assignment determines the level of scrutiny the device is subjected to for conformity assessment. Class A devices offer the lowest risk, Class B low to moderate hazard, Class C moderate to high risk, and Class D the highest risk. The level of regulatory oversight and the evidence required to support the product’s conformity assessment to Essential Principles for Safety and Performance becomes more robust and demanding as the device’s classification increases from A to D (21,22). Medical devices other than condoms and gloves are not quality assured and marketed without undergoing a process of evaluation to determine the product’s quality, safety, and performance. There is no assurance that only medical devices the NRA has duly authorised are allowed to be manufactured, imported, distributed, sold or supplied to end users.

### VL

The vigilance regulatory function had a quality score of 27% due to the lack of legal provisions, regulations, and guidelines required to define the regulatory framework of vigilance, leaving the Zimbabwean population at risk of medical devices that do not continue to meet requirements for quality, safety and performance throughout the product’s lifecycle. An established, implemented and maintained reporting system must be established to monitor medical device quality, safety and performance. The regulatory function is essential because even mature regulatory jurisdictions such as USFDA and the EU were not spared from adverse incidents and recalls of medical devices, which called for an overhaul of the regulatory system in the EU context (8). These deficiencies imply that the magnitude of adverse events related to medical devices is unknown in the Zimbabwean context. The human and economic costs of these deficiencies can be severe. Legislation must mandate the economic operators (manufacturers, importers, distributors, authorised representatives and wholesalers) to collect and evaluate the experiences gained from medical devices placed on the market. The economic operators must record and investigate any incident report they receive, implement field safety corrective action proportional to the identified deficiency and inform the regulatory authority when the law demands so. Regulators must be informed when there is a severe public health threat or when death or serious deterioration in health occurred or may have occurred to the user, patient or another person. The incident may be linked to the medical device’s quality, safety or performance. The regulatory authority may monitor the manufacturer’s investigation to determine the root cause (4,5).

### MC

An effort has been made to establish import and export regulations for medical devices attaining a 40% score. However, there are no legal provisions concerning import activities, including permanent regulatory intervention at designated entry and exit ports where medical devices are moved. Similarly, there were no legal provisions to authorise market surveillance and control activities, including medical device sampling from different supply chain points. These deficiencies expose the country to SF medical devices and may compromise:

- patients’ safety and health outcomes,
- national economy due to the disease burden,
- public trust in the healthcare system, and
- the international fight against serious health challenges such as malaria and antimicrobial resistance (23).

Additionally, the lack of legal provisions and regulations addressing the role of NRA in dealing with SF medical products makes it challenging to implement this regulatory function at the present moment.

There is a shortage in the literature regarding SF medical products in Africa and the Middle East. Studies have been mainly on medicines; this clearly shows that more needs to be done regarding SF medical devices and must be prioritised to ensure acceptable quality, safety and performance of medical devices (23). Challenges in managing medical products have been documented in the SADC region; however, more needs to be known regarding medical devices, including IVDs. A study conducted to map existing frameworks, mechanisms and approaches to prevention, detection and response (PDR) to SF medical products concluded that most countries had the legal mandate to implement measures to curb SF medical products. Still, they were not doing so (14). Product failures have been reported in the United States (US) and European Union, resulting in regulatory regime reforms. The US and EU regulations are highly esteemed. However, high incidences of clinical adverse events, high recall rates, and frequent phase-out of some medical devices have resulted in overhauling the regulations (8). The problem may be more for a country like Zimbabwe that needs legislation to curb SF medical devices.

### LI

There were no legal provisions, regulations, or guidelines for licensing economic operators along the value chain of medical devices resulting in an overall score of 62%. The score implies that economic operators involved in the medical devices supply chain are not licensed using a licensing regime that is proportional to the role in the medical device’s value chain and the medical device’s risk classification. Consequently, there is no traceability of medical devices throughout the supply chain of medical devices, except for condoms and gloves. The NRA must be empowered by legislation to issue licenses and suspend or revoke establishment permits. In cases of post-licensing changes, the legislation or regulations must require the economic operators to notify the regulatory authority.

### LT

The laboratory testing regulatory function had the highest score (88%) due to the lab testing conducted for condoms and gloves, excluding other medical devices, including IVDs. This score may be due to assessments by external bodies since the laboratory is ISO 17025 accredited. However, due to the wide range and complexities of medical devices, medical devices other than condoms and gloves are not independently verified by laboratory testing where applicable based on the risk class of the medical device. The non-verification for market authorisation, vigilance, market surveillance and control or post-authorisation changes exposes the population to the risk of SF medical devices. Due to the wide range of medical devices and the associated laboratory expertise, the NRA may outsource part or most laboratory testing. The details of the outsourced testing services’ responsibilities, duties and roles in these structures should be clearly defined and documented, including their accreditation status. The NRA must implement mechanisms to select, monitor and evaluate laboratories performing laboratory work before fulfilling the regulatory requirements. The evaluation may be done through onsite audits using ISO 15189 and ISO 17025 requirements to assess policies and procedures for the tests conducted in individual laboratories. Having an ISO accreditation is a good starting point. However, more is needed, as anecdotal evidence has shown that labs that may have ISO 15189 accreditation failed the audit conducted by the WHO for the labs to be listed as WHO Performance Evaluation Laboratories (PEL) for IVDs.

Performance evaluation is critical to verify a manufacturer’s claims for a medical device. Therefore, it is of paramount importance for the NRA to conduct an assessment and analysis of data to establish or verify the scientific validity, the analytical and, where applicable, the clinical performance of a medical device to ensure that the intended purpose and the classification is appropriate based on the specific disorder, condition or risk factor of interest that it is intended to detect, define or differentiate(4,5).

### RI

The regulatory inspection had an implementation quality score of 68% due to market authorisation inspections conducted for establishments involved in the supply chain of condoms and gloves. However, legal provisions, regulations and guidelines for regulatory inspection were not explicit for medical devices. Establishments involved in other medical devices besides condoms and gloves are unknown regarding compliance with perceived standards. The NRA must publish the list of measures that all the economic operators must comply with, and these standards must be the basis upon which the economic operators are audited for compliance. Regulatory inspection activities must be supported by a comprehensive set of legal provisions, regulations and guidelines which provide the necessary mandate to implement all activities related to this regulatory function.

### CT Oversight

The CT oversight regulatory scored the least (18%). The MASCA mentions clinical trials in the context of medicines only, but not for medical devices. Some classes of medical devices require clinical trials to be conducted to support registration and market authorisation of these products. Before registration and market authorisation, the products are classified as IMPs. The NRA needs to oversee these products before, during and after the implementation of CTs. The NRA must have the legal mandate to authorise, regulate and, if necessary, terminate CTs in line with the international medical research ethics guidelines and principles. Before the trial commences, the NRA should have trained and competent staff to review CT protocols, also reviewed by an Independent Ethics Committee (IEC). The IEC should review the protocols and should have the authority, when necessary, to require protocol revisions. The CT review committee should comprise members that are competent and skilled and ensure that there is no conflict of interest. The finding in this study is that there is CT oversight by the NRA for investigational medical devices undergoing CT in Zimbabwe due to the lack of the following:

- Legal provisions and regulations for CTs oversight.
- Legal provisions and regulations require research centres, researchers, sponsors, clinical research organisations (CROs) and all relevant institutions in the clinical trial to comply with good clinical practice (GCP).
- Legal provisions and regulations stipulate that authorisation from NRA and notification to the NRA are required where there are changes to the original protocol or any relevant documents of the CT.
- Legal provisions, regulations and guidelines require that IMPs comply with good manufacturing practice (GMP) for IMPs.
- Legal provisions or regulations cover circumstances where the routine CT evaluation and procedure may not be followed (e.g. for public-health interests).
- Legal provisions, regulations or guidelines exist for NRA to inspect, suspend or stop CTs.

These deficiencies subject the Zimbabwean population to products needing acceptable quality, safety and performance during CTs.

Furthermore, the NRA is unaware of the CTs conducted and those in progress; the research participants’ rights are not guaranteed because the participants are uninformed. This is unacceptable today, considering the previous well-documented violations: “*Tuskegee Syphilis Study from 1932 to 1972, Nazi medical experimentation in the 1930s and 1940s, and research conducted at the Willowbrook State School in the 1950s and 1960s. As the aftermath of these practices, wherein uninformed and unaware patients were exposed to a disease or subject to other unproven treatments, became known, the need for rules governing the design and implementation of human-subject research protocols became very evident.”* (24).

Cross-cutting indicators were generally in place; policy and strategic planning, leadership and crisis management, transparency, accountability and communication, quality and risk management systems, regulatory process, resources (Human, financial, infrastructure, equipment and information management systems), monitoring progress and assessing impact. However, the indicators were sufficient for regulating condoms and gloves only. There were some vacant staff positions compared to the current establishment of the NRA. The competence framework was sufficient for the existing scope of products. There is a need to review the operational and competency framework to meet the scope and complexity of other medical devices. The competency framework must include the following aspects:

- Scientific and Health Concepts: Understanding and applying “evolving basic and translational science, regulatory science and public health concepts to drive new approaches to improve healthcare product development, review and oversight” (25)
- Ethics: Ability to integrate and demonstrate core values, integrity and accountability.
- Business Acumen: Ability to successfully leverage systems and processes to operate a regulatory function.
- Communication: Ability to convey or exchange information with stakeholders within and outside the organisation.
- “Leadership: Ability to direct and contribute to initiatives within the organisation, with groups engaged in developing good regulatory practice and policy, and within the regulatory profession.
- Regulatory Frameworks and Strategy: Knowledge of regulatory frameworks and external environments and the ability to apply these to regulatory solutions throughout the product lifecycle.
- Product Development and Registration: Knowledge of the research and development, preclinical and clinical steps and related regulations in healthcare product development.
- Postapproval/Post-market: Knowledge of requirements and processes for maintaining a product on the market, reporting and surveillance”(25,26)

The NRA needs to improve mechanisms to promote transparency, accountability and communication by publishing summary technical evaluation reports for approved registration MA, inspections, and rejected applications.

### Study Limitations

The study was conducted in the context of MCAZ as the NRA. However, the Medical Laboratory and Clinical Scientists Council of Zimbabwe and the National Microbiology Reference Laboratory were not benchmarked. The two institutions register IVD Medical for priority pathogens. The registration is used as an eligibility criterion for products to be procured through the national tendering process.

## Conclusions

The readiness of the NRA in Zimbabwe to regulate medical devices is below optimum as it is at the lowest possible score that can be attained using the GBT+ medical devices methodology. The NRA does not have an explicit legal mandate to regulate medical devices. The MASCA needs more clarity on legal provisions for medical devices regulatory system framework, registration and market authorisation, vigilance, market surveillance control, licensing establishment, regulatory inspection, laboratory testing and clinical trials. The legal provisions, regulations, and guidelines are inadequate for effectively regulating medical devices. The medical devices regulation requires review for it to be robust and fit-for-purpose, responsive, oriented to the outcome, predictable based on a standard and transparent approach, and the level of scrutiny proportionate to the risk classification of the medical device.

## Data Availability

All relevant data are within the manuscript and its Supporting Information files.

## Acknowledgements

The authors would like to acknowledge the cooperation of the staff at NRA IN Zimbabwe for their assistance during the interviews and availing the evidence for the assessment to be successiful.

